# Extreme γ’ fibrinogen levels in COVID-19 patients

**DOI:** 10.1101/2022.01.20.22269321

**Authors:** David H. Farrell, Matthew Hudkins, Heather Hamilton, Samantha J. Underwood, Elizabeth N. Dewey, Diana E. Kazmierczak, Steven C. Kazmierczak, William B. Messer, Akram Khan, Martin A. Schreiber

## Abstract

**Background:** COVID-19 progression can be accompanied by a “cytokine storm” that leads to secondary sequelae such as thrombosis and acute respiratory distress syndrome. Several inflammatory cytokines have been associated with COVID-19 progression, but have far too much daily intra-individual variability to be useful in tracking the course of the disease. In contrast, we have shown that the inflammatory biomarker γ’ fibrinogen (γ’ Fbg) has a 6-fold lower coefficient of variability compared to other inflammatory markers such as hs-CRP. *Objectives:* The aims of the study were to measure γ’ Fbg in serial blood samples from COVID-19 patients at a tertiary care medical center in order to investigate its association with clinical measures of disease progression.

**Methods:** COVID-19 patients at a tertiary care medical center were retrospectively enrolled between 3/16/2020 and 8/1/2020. γ’ Fbg was measured using a commercial ELISA. *Results:* Our results showed that nine out of the seventeen patients with COVID-19 had extremely high levels of γ’ Fbg. γ’ Fbg levels were significantly associated with the need for ECMO and with mortality.

**Conclusions:** We found that COVID-19 patients can develop extraordinarily high levels of γ’ Fbg. The previous highest γ’ Fbg level that we are aware of was 80.3 mg/dL found in a study of 10,601 participants in the ARIC study. These results have several important clinical implications. γ’ Fbg contains a high affinity binding site for thrombin that binds to anion-binding exosite II on thrombin and protects it from inactivation by heparin. High levels of γ’ Fbg therefore provide a reservoir of heparin-resistant clot-bound thrombin when the γ’ Fbg is clotted. These findings have potential implications regarding prophylactic anticoagulation of COVID-19 patients and suggest that heparin prophylaxis may be less effective than using other anticoagulants, particularly direct thrombin inhibitors.

## 1. Introduction

Severe acute respiratory syndrome coronavirus (SARS-CoV-2) causing coronavirus disease 2019 (COVID-19) has infected more than 322 million individuals worldwide and caused more than 5.5 million deaths [1]. COVID-19 progression can be accompanied by a “cytokine storm” [2] that leads to secondary sequelae such as thrombosis [3, 4] and acute respiratory distress syndrome (ARDS) [5]. Several inflammatory cytokines, including high-sensitivity C-reactive protein (hs-CRP) and interleukin-6 (IL-6), have been associated with COVID-19 progression [6], but have far too much daily intra-individual variability to be useful in tracking the course of the disease. In contrast, we have shown that the inflammatory biomarker gamma prime fibrinogen (γ’ Fbg) has a 6-fold lower coefficient of variability compared to other inflammatory markers such as hs-CRP [7]. The mRNA for the γ chain of γ’ Fbg, expressed from the *FGG* gene, is upregulated 8.3-fold by IL-6 *in vitro* [8], so that γ’ fibrinogen is likely to be a more stable and therefore superior surrogate marker to IL-6 as well.

γ’ Fbg arises from alternative mRNA processing [9] that results in the substitution of the carboxyl terminal four amino acids with a different twenty-amino acid sequence. The γ’ chain pairs with the more common γA chain to form γA/γ’ fibrinogen, whereas the majority of fibrinogen is γA/γA fibrinogen. γ’ fibrinogen typically constitutes ∽7% of total fibrinogen in plasma [10], although this varies widely among individuals [11, 12, 13, 14]. The normal range of total fibrinogen is 150-300 mg/dL, whereas γ’ Fbg has a wide reference interval in healthy individuals from 8.8-55.1 mg/dL [13].

γ’ fibrinogen has several biochemical and biophysical properties that distinguish it from the more common γA isoform. Clots made from γ’ fibrinogen in the presence of coagulation factor XIII (a.k.a. “fibrin-stabilizing factor” in earlier nomenclature) are highly resistant to fibrinolysis [15, 16, 17,18]. In addition, the γ’ chain contains a high affinity binding site for thrombin [19, 20, 21, 22], and clots made from γ’ fibrinogen have an altered clot architecture [17, 18, 23, 24]. Importantly, thrombin that is bound to the γ’ chain remains bound in the growing fibrin clot and is resistant to inactivation by antithrombin and heparin [25]. This is due to the fact that the γ’ chain binds to the heparin binding site on thrombin[20]. Because of this, we measured γ’ Fbg in serial blood samples from COVID-19 patients at a tertiary care center in order to investigate its association with clinical measures of disease progression.

## 2. Materials and Methods

### 2.1 Patients

Hospitalized COVID-19 patients at Oregon Health & Science University (OHSU) Hospital were retrospectively enrolled between 3/16/2020 and 8/1/2020. Samples were collected earlier and de-identified, and stored frozen in a biological repository. This study was approved by the OHSU IRB, STUDY00021409, and was carried out in accordance with The Code of Ethics of the World Medical Association (Declaration of Helsinki). Patients who had at least two EDTA plasma draws were randomly selected from this time frame.

Data from the Atherosclerosis Risk In Communities (ARIC) study were used to compare the distribution of γ’ Fbg to these historical controls [10]. For this purpose, we extracted 10,601 ARIC participants who had their γ’ Fbg levels measured. In the ARIC study, γ’ Fbg was measured in plasma samples, and were collected from 1993-1995. Participants with prevalent cardiovascular disease (coronary heart disease, heart failure, peripheral artery disease, or stroke) were excluded. It should be noted that γ’ Fbg is stable for years when stored frozen at -80°C, as shown by the similar mean and median values in several previously published studies [10, 12, 13].

### 2.2 γ’ Fbg assays

γ’ Fbg was measured using the GammaCoeur ELISA according to the manufacturer’s instructions (Zeus Scientific, Branchburg NJ/Gamma Diagnostics, Portland, OR; patent pending) and measured at 450 nm in a BioTek ELx808 (BioTek Instruments, Winooski, VT).

### 2.3 Statistical analysis

There were 17 patients in the COVID-19 cohort with multiple lab values recorded throughout their hospital stay. The number of measurements ranged from 1-5 for each patient, with one outlier patient having 67 reported total fibrinogen levels. To simplify analysis, patient summary data were calculated. For each patient, the minimum, median, and maximum lab values were calculated and used for analysis. We did not calculate the mean, because the distribution for γ’ Fbg is not normally distributed. We also evaluated the minimum and the maximum, because γ’ Fbg levels in this cohort were much higher than previously reported [10]. Standard descriptive statistics were used to summarize patient demographic and clinical characteristics. The median γ’ Fbg in COVID-19 patients was compared to the median γ’ Fbg in the ARIC study [10] using z-test. Histograms were constructed to visually compare the distribution of γ’ Fbg in both cohorts. Within the COVID-19 cohort, γ’ Fbg was evaluated for associations with ARDS and ECMO using Wilcoxon Rank-Sum tests, and Holm’s correction for multiple comparisons was used to identify significant associations. Analysis was conducted in JMP 15.2 and SAS 9.4 (SAS Inc., Cary, NC for both). Significance was assessed at *p*<0.05.

## 3. Results

The γ’ Fbg levels in the COVID-19 patients were compared to historical controls (Fig. 1) from ARIC [10]. The median γ’ Fbg levels in ARIC were 29.6 mg/dL (IQR=24.5-35.7 mg/dL) compared to 42.1 mg/dL (IQR=8.7-70.8 mg/dL) in the COVID-19 patients. The highest γ’ Fbg level in the ARIC study was 80.3 mg/dL [10]. In the COVID-19 cohort, the highest γ’ Fbg level was 261 mg/dL. Descriptive statistics for the distribution of γ’ Fbg as well as D-dimer and total fibrinogen are reported in Table 2. Descriptive statistics for patients from the ARIC study are not reported here, as they are well documented [10].

**Fig. 1.**
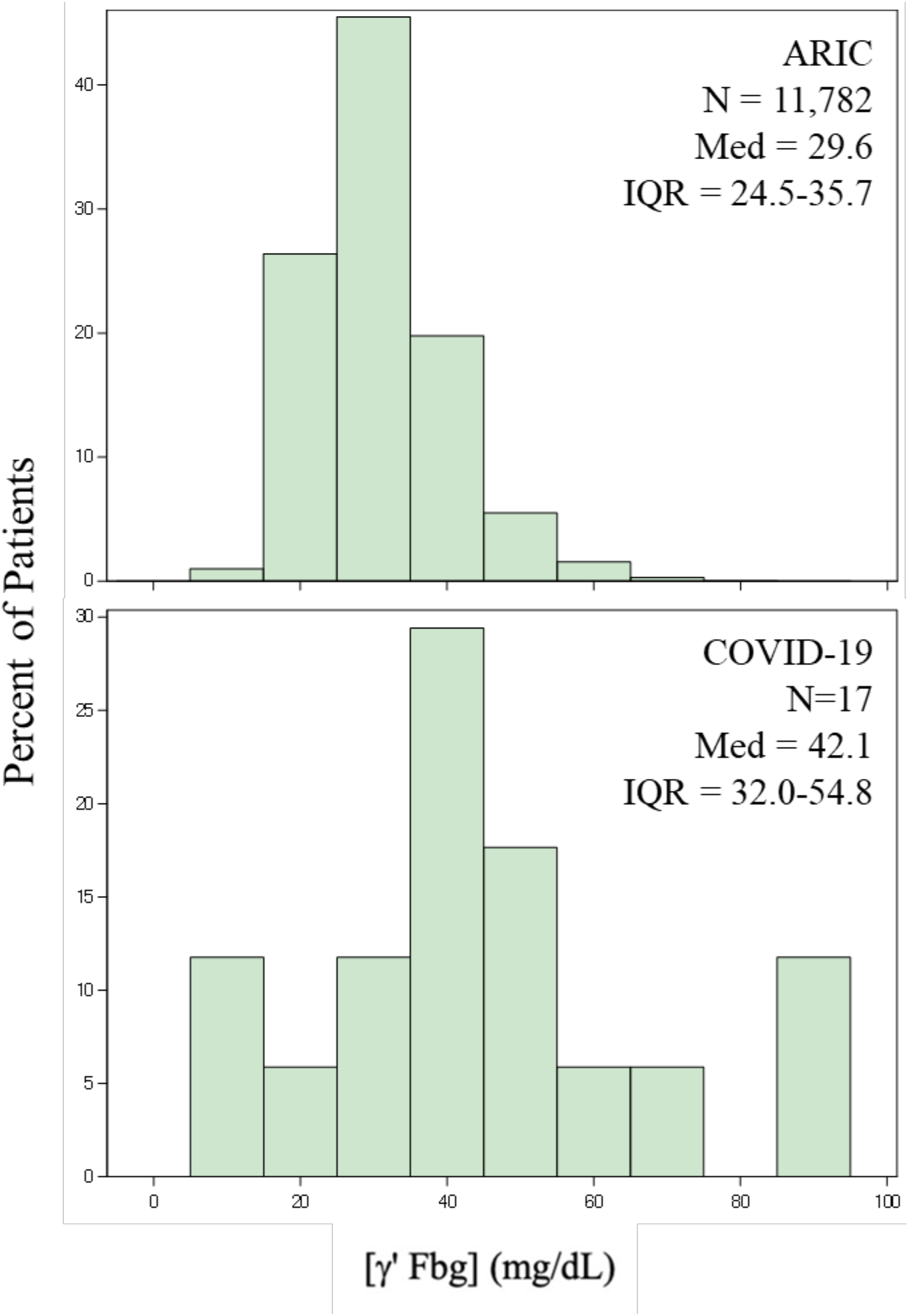
Comparison Between γ’ Fbg Levels in the ARIC Cohort and COVID-19 Patients. The median value for γ’ Fbg is significantly higher in the COVID-19 cohort (p<0.01).

**Table 1.** Patient Characteristics for Hospitalized COVID-19 patients at Oregon Health & Science University (OHSU) Hospital Admitted between 3/16/2020 and 8/1/2020.

| Patient Characteristic | N | % or Median (IQR) |
| --- | --- | --- |
| Age | 17 | 46 (36, 50) |
| LOS | 17 | 18.2 (14, 26.7) |
| Gender (male) | 11 | 64.7 |
| ARDS | 9 | 52.9 |
| ECMO | 6 | 35.3 |
| Days on ECMO | 6 | 12.9 (8.9, 31.7) |
| VTE | 5 | 29.4 |
| Intubated | 10 | 58.8 |
| Re-intubated | 4 | 40.0 |
| Days intubated | 10 | 5.5 (1.3, 6.1) |
| COVID Treatments |  |  |
| Remdesivir | 7 | 41.2 |
| Sarilumab | 1 | 5.9 |
| Dexamethasone | 6 | 35.3 |

**Table 2.** Distribution of D-Dimer, γ’ Fbg, and Total Fibrinogen in COVID Patients.

| Lab Value* | Min | 25th %ile | Median | Mean | 75th %ile | Max |
| --- | --- | --- | --- | --- | --- | --- |
| *D-Dimer (number of measurements range 1-6 per hospital stay) (mg/L)* |  |  |  |  |  |  |
| Median D-Dimer | 1.00 | 1.39 | 1.80 | 1.80 | 1.80 | 2.68 |
| Min D-Dimer | 0.27 | 0.31 | 0.31 | 0.31 | 0.31 | 0.31 |
| Max D-Dimer | 4 | 4 | 4 | 4 | 4 | 4 |
| *$\gamma'$ Fbg (number of measurements range 1-6 per hospital stay) (mg/dL)* |  |  |  |  |  |  |
| Median $\gamma'$ Fbg | 23.58 | 28.69 | 42.06 | 49.63 | 70.80 | 81.38 |
| Min $\gamma'$ Fbg | 5.50 | 5.50 | 11.55 | 14.50 | 22.50 | 28.70 |
| Max $\gamma'$ Fbg | 85.99 | 85.99 | 88.25 | 88.25 | 136.91 | 150 |
| *Total Fibrinogen (number of measurements range 1-5 per hospital stay) (mg/dL)\*\** |  |  |  |  |  |  |
| Median Total Fibrinogen | 343 | 418 | 488 | 488 | 488 | 488 |
| Min Total Fibrinogen | 330 | 340 | 350 | 390 | 450 | 460 |
| Max Total Fibrinogen | 750 | 750 | 750 | 750 | 750 | 750 |

\*Median, Min, and Max Lab Values are calculated from patient's daily measurements; e.g., the statistics for the minimum is based on the minimum observed over the course of the hospital stay for each of the 17 patients in the COVID cohort.
\*\*one outlier patient had 67 measurements of total fibrinogen.

There were several notable shifts in γ’ Fbg in patients who developed ARDS, were placed on ECMO, or died in-hospital (Table 3). Patients who developed ARDS reported higher peak levels of γ’ Fbg (max = 88.8 *vs*. 68.3 mg/dL, p = 0.53). Patients who progressed to ECMO reported significantly higher minimum levels for γ’ Fbg (min = 38.5 *vs*. 17.4 mg/dL, p = 0.01). The two patients who died reported significantly higher minimum and median γ’ Fbg levels compared to the patients who lived (min = 86 *vs*. 22.9 mg/dL, p = 0.03; median = 87.8 *vs*. 40.3 mg/dL, p = 0.03). There were no associations between elevated γ’ Fbg levels and VTE events, and patient summary data were similar for the minimum and median γ’ Fbg levels. Patients who experienced a VTE event had slightly lower peak γ’ Fbg (max = 71.5 *vs*. 85.1 mg/dL, p = 0.87). However, it should be noted that the small number of deaths limits the conclusions that can be drawn from the statistical comparison.

**Table 3.** γ’ Fbg Levels by In-Hospital Outcomes.

| Outcome | Median (IQR) for Patient Summary Data of $\gamma'$ Fbg | | | |
| --- | --- | --- | --- | --- |
|  | N | Patient Min | Patient Median | Patient Max |
| *ECMO* |  |  |  |  |
| No | 11 | 17.4 (8.3, 29.7) | 40.1 (22.5, 50.2) | 70.8 (50.5, 136.9) |
| Yes | 6 | 38.5 (28.8, 86) | 55.9 (40.3, 87.4) | 85.1 (71.5, 90.5) |
| Unadjusted p-value |  | 0.01 | 0.17 | 0.77 |
| *ARDS* |  |  |  |  |
| No | 8 | 16.3 (7.3, 30.1) | 36.0 (17.8, 43.1) | 68.3 (45.4, 131.8) |
| Yes | 9 | 28.8 (21.2, 42.6) | 51.8 (40.3, 60.1) | 88.8 (71.5, 90.5) |
| Unadjusted p-value |  | 0.12 | 0.54 | 0.53 |
| *Discharge Disposition* |  |  |  |  |
| Expired | 2 | 86 (86, 86) | 87.8 (87.4, 88.2) | 89.7 (88.8, 90.5) |
| Home | 15 | 22.9 (9.1, 30.4) | 40.3 (31.9, 51.8) | 71.5 (50.5, 126.7) |
| Unadjusted p-value |  | 0.03 | 0.03 | 0.55 |
| *In-Hospital VTE Event* |  |  |  |  |
| No | 12 | 20.5 (8.7, 34.6) | 43.1 (27.2, 55.9) | 85.1 (63.3, 124.7) |
| Yes | 5 | 28.8 (22.9, 29.7) | 40.3 (40.1, 54.8) | 71.5 (50.5, 90.5) |
| Unadjusted p-value |  | 0.43 | 0.67 | 0.87 |

All 17 patients had more than one measured lab value during their hospital stay; analysis was conducted on patient summary data. Values reported here represent the median of the patient's calculated lab value.
“Patient Median” is the median $\gamma'$ Fbg calculated from all patient's median $\gamma'$ Fbg levels.
“Patient Min” is the median $\gamma'$ Fbg calculated from all patient's minimum $\gamma'$ Fbg levels.
“Patient Max” is the median $\gamma'$ Fbg calculated from all patient's maximum $\gamma'$ Fbg levels.
All reported p-values are unadjusted, and were calculated using Wilcoxon Rank-Sum test; no comparison was statistically significant after adjusting for multiple comparisons using Holm's correction.

Two longitudinal case studies were also informative. One patient was a male who was admitted to the hospital following a COVID-19 outbreak in his care facility and was suspected of having SARS-CoV-2. The diagnosis was confirmed upon admission. Chest X-rays revealed no focal consolidation on admission, but by day 5, worsening opacities were observed. On day 8, his clinical condition declined, with increased oxygen needs and hypoxia. The patient eventually recovered and was discharged on day 18. When the patient’s γ’ Fbg levels were measured retrospectively, a significant increase in γ’ Fbg levels was observed on day 8 (Fig. 2). There was thus a temporal association with the worsening of clinical symptoms and γ’ Fbg levels.

**Fig. 2.**
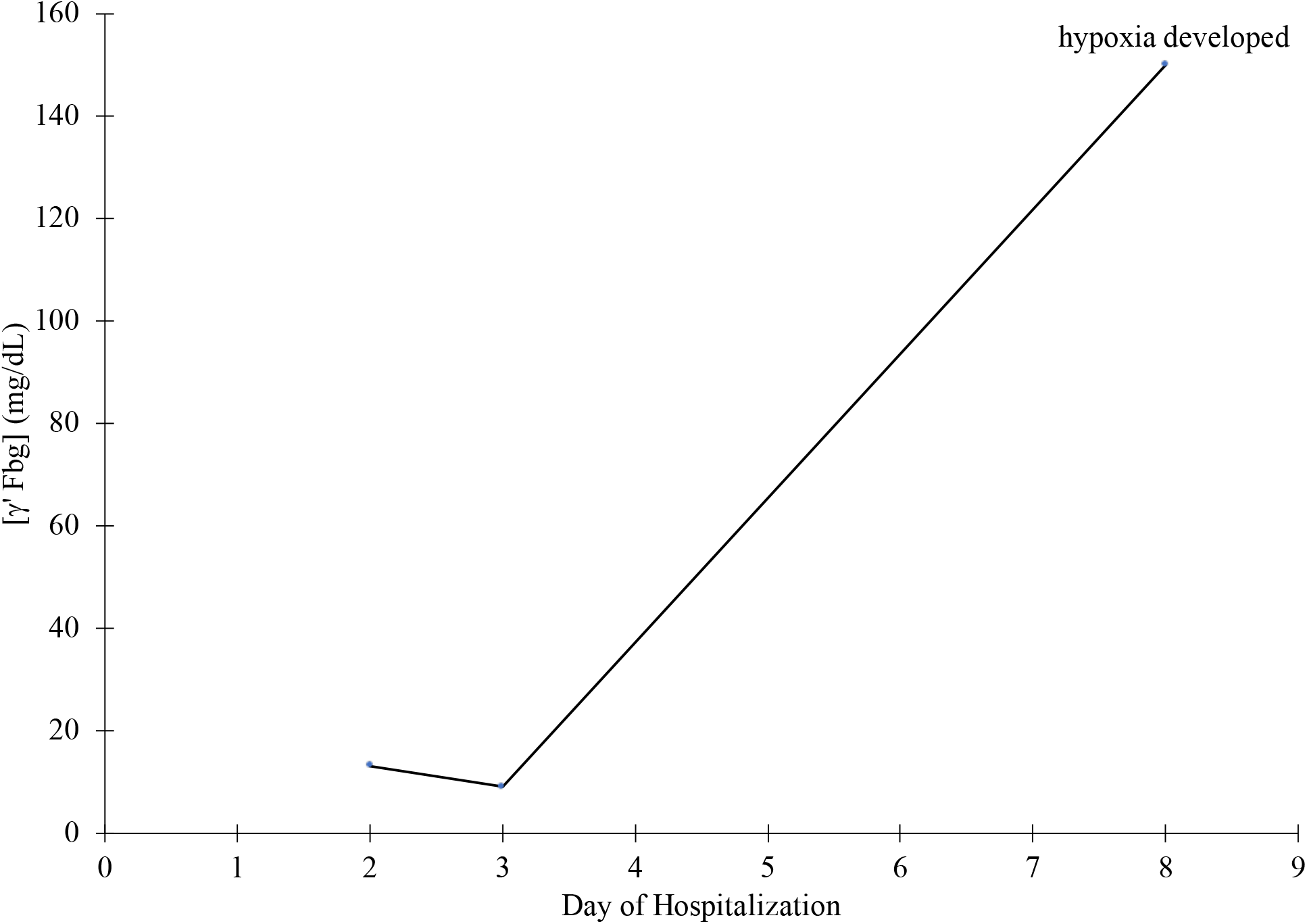
Association between γ’ Fbg levels and the development of hypoxia in one patient.

The second patient was a male admitted to the emergency department 10 days after symptom onset, which included COVID-19 pneumonia, fever and cough. At admission, the patient had respiratory failure requiring high flow ventilation, which subsequently stabilized. Eight doses of methylprednisone, 40 mg q12, were given based on clinical symptoms, starting on day 2. The patient subsequently improved and was discharged on day 10. When the patient’s γ’ Fbg levels were measured retrospectively, the peak of γ’ Fbg coincided with the worsening symptoms, and subsequently decreased after the administration of methylprednisone (Fig. 3). (It should be noted that this was before the widespread use of dexamethasone.) To our knowledge, this patient had the highest level of γ’ Fbg ever reported, 260 mg/dL, which was 23.1% of the total fibrinogen level of 1,121 mg/dL. For comparison, γ’ Fbg levels average 7% of total fibrinogen [10], although γ’ Fbg has a wide reference interval in healthy individuals from 0.088-0.551 mg/dL.

**Fig. 3.**
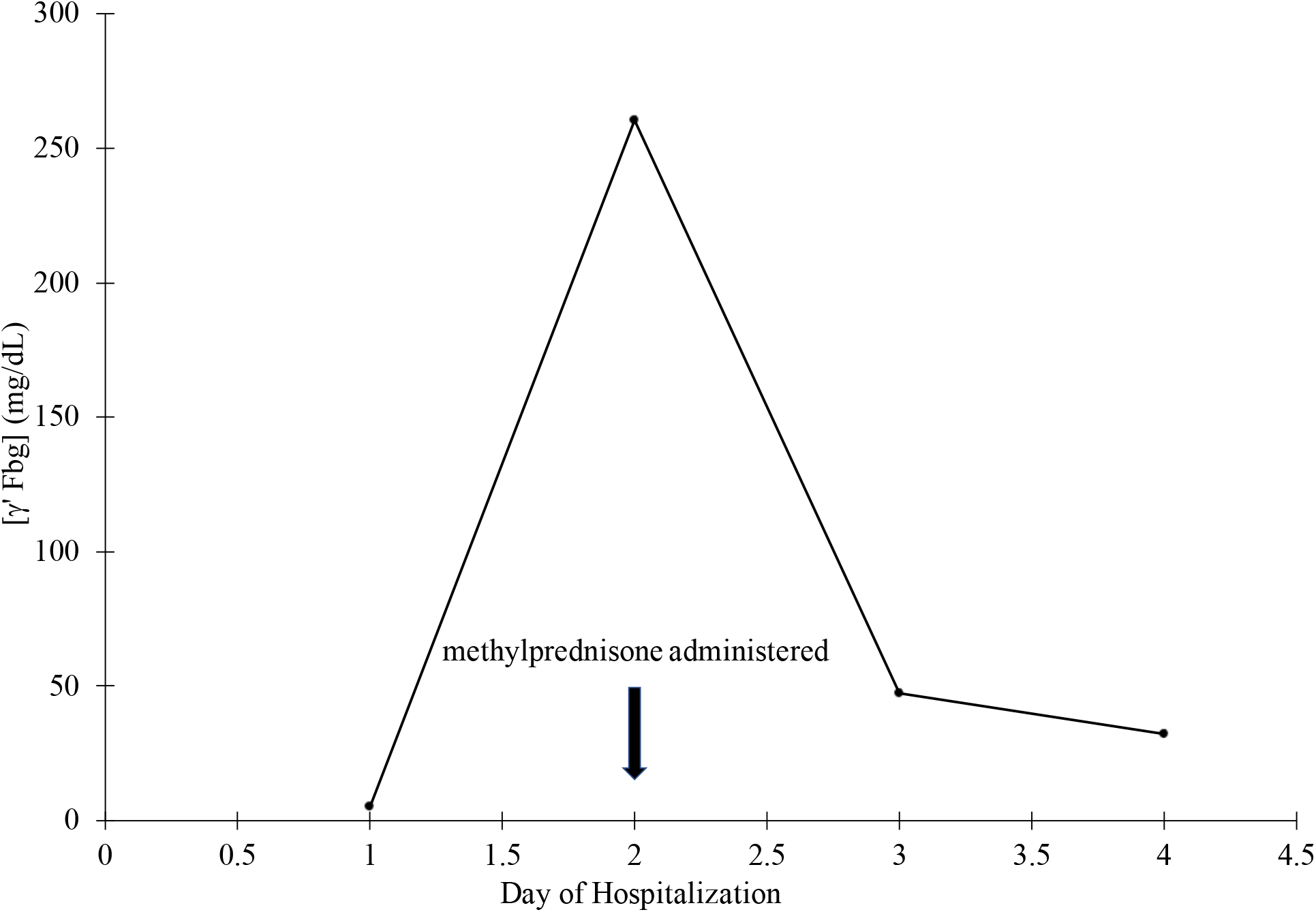
Association between γ’ Fbg levels and steroid treatment in one patient.

## 4. Discussion

The finding that COVID-19 patients can develop extraordinarily high levels of γ’ Fbg has several important clinical implications. γ’ Fbg contains a high affinity binding site for thrombin [19, 20, 21, 22]. This binds to anion-binding exosite II on thrombin [20] and protects it from inactivation by heparin [25]. The mechanism behind the heparin resistance stems from the fact that unfractionated heparin also binds to exosite II; therefore, heparin cannot bind to thrombin that is bound to γ’ Fbg. High levels of γ’ Fbg therefore provide a reservoir of heparin-resistant clot-bound thrombin when the γ’ Fbg is clotted (Fig. 4).

**Fig 4.**
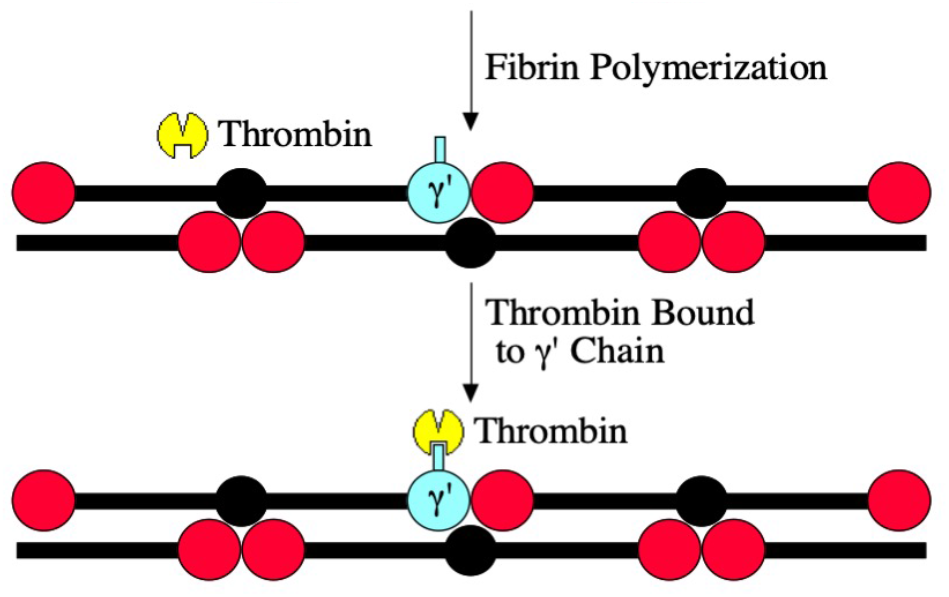
Model of γ’ Fbg mechanism of thrombosis. The model starts with free thrombin cleaving fibrinopeptides A and B from all forms of fibrinogen *via* its active site (cutout triangle). This converts the fibrinogen to fibrin, which polymerizes into an insoluble clot. The free thrombin can then bind to γ’ fibrin *via* thrombin anion-binding exosite II (cutout square), concentrating active thrombin on the growing fibrin clot. This bound thrombin is resistant to heparin, since heparin binds to anion-binding exosite II, which is blocked by the γ’ chain.

The rapid decrease in γ’ Fbg levels following the administration of methylprednisone (Fig. 3) was unexpected, given the reported half-life of fibrinogen antigen of 88 hours [26]. However, there are some possibilities that could explain the rapid loss of γ’ Fbg seen in this patient. One is that there was ongoing consumption of γ’ Fbg due to intravascular coagulation. The association of elevated D-dimer levels in COVID-19 patients is consistent with this hypothesis, since elevated D-dimers are indicative of fibrin formation followed by fibrinolysis [3]. Another possible explanation is that γ’ Fbg may have a shorter half-life than unfractionated fibrinogen, since the half-life of γ’ Fbg has never been measured experimentally. However, this would imply that γ’ Fbg has a different clearance mechanism than unfractionated fibrinogen; there is no evidence for this. Therefore, the mechanism(s) for the rapid decrease in γ’ Fbg levels remains unclear.

These findings have potential clinical implications regarding prophylactic anticoagulation of COVID-19 patients. The resistance of γ’ Fbg-bound thrombin to heparin suggests that heparin prophylaxis may be less effective than treatment with other anticoagulants, particularly direct thrombin inhibitors. It is possible that inhibition of factor Xa by current DOACs may also reduce the levels of active thrombin and thereby prevent activation of thrombin substrates by γ’ Fbg-bound thrombin, including factor V, factor VIII, factor XI, factor XIII, and fibrinogen, as well as platelet substrates such as PAR-1 and PAR-4. In addition, it is also possible that warfarin anticoagulation may be effective at preventing thrombosis due to γ’ Fbg-bound thrombin by reducing the levels of active vitamin K-dependent coagulation factors, and therefore, generation of thrombin.

## 5. Conclusions

In summary, we suggest that γ’ Fbg levels may be an important marker of disease progression in patients with COVID-19, and γ’ Fbg may play a role in the coagulation abnormalities noted in these patients.

## Data Availability

All data produced in the present study are available upon reasonable request to the authors

## Abbreviations

γ’ Fbg: γ’ fibrinogen
ARDS: acute respiratory distress syndrome
ARIC: Atherosclerosis Risk In Communities study
COVID-19: coronavirus disease 2019
DOAC: direct oral anticoagulant
ECMO: extracorporeal membrane oxygenation
ELISA: enzyme-linked immunosorbent assay
hs-CRP: high-sensitivity C-reactive protein
IL-6: interleukin-6
IRB: institutional review board
OHSU: Oregon Health & Science University
PAR-1: protease-activated receptor-1
PAR-4: protease-activated receptor-4
Sars-CoV-2: severe acute respiratory syndrome coronavirus

## Funding and acknowledgments

This research did not receive any specific grant from funding agencies in the public, commercial, or not-for-profit sectors.

---

**Supplementary Table 1..** Raw data for each COVID-19 patient.

| Age | Gender | Day in Hospital | [γ' Fbg] (mg/dL) | ARDS | ECMO | Death |
| --- | --- | --- | --- | --- | --- | --- |
| 36-40 | M | 9 | 88.9 | Yes | Yes | No |
|  |  | 10 | 25.4 |  |  |  |
|  |  | 13 | 68.0 |  |  |  |
|  |  | 14 | 38.0 |  |  |  |
|  |  | 15 | 32.7 |  |  |  |
|  |  | 16 | 17.4 |  |  |  |
| 46-50 | F | 1 | 8.34 | No | No | No |
|  |  | 3 | 49.2 |  |  |  |
|  |  | 4 | 65.8 |  |  |  |
|  |  | 7 | 22.5 |  |  |  |
|  |  | 9 | 17.0 |  |  |  |
| 71-75 | M | 2 | 13.1 | No | No | No |
|  |  | 3 | 9.12 |  |  |  |
|  |  | 8 | >150 |  |  |  |
| 46-50 | M | 2 | 6.82 | No | No | No |
|  |  | 3 | 6.17 |  |  |  |
|  |  | 4 | 28.7 |  |  |  |
|  |  | 5 | 16.3 |  |  |  |
| 46-50 | F | 2 | 71.5 | Yes | Yes | No |
|  |  | 4 | 39.6 |  |  |  |
|  |  | 6 | 28.8 |  |  |  |
|  |  | 7 | 40.3 |  |  |  |
|  |  | 14 | 63.9 |  |  |  |
| 51-55* | M | 1 | 5.23 |  |  |  |
|  |  | 2 | 260.4 |  |  |  |
|  |  | 3 | 47.1 |  |  |  |
|  |  | 4 | 32.2 |  |  |  |
| 21-25 | F | 1 | 44.1 | No | No | No |
|  |  | 2 | 70.8 |  |  |  |
|  |  | 3 | 34.8 |  |  |  |
| 31-35 | F | 4 | 122.3 | Yes | Yes | No |
|  |  | 5 | 48.8 |  |  |  |
|  |  | 6 | 42.6 |  |  |  |
|  |  | 9 | 60.1 |  |  |  |
|  |  | 11 | 72.4 |  |  |  |
| 56-60 | M | 1 | 21.4 | Yes | No | No |
|  |  | 2 | 79.9 |  |  |  |
|  |  | 4 | 89.9 |  |  |  |
|  |  | 5 | 29.7 |  |  |  |
|  |  | 6 | 21.2 |  |  |  |
| 21-25 | F | 4 | 22.9 | Yes | Yes | No |
|  |  | 6 | 33.3 |  |  |  |
|  |  | 7 | 30.6 |  |  |  |
|  |  | 8 | 28.7 |  |  |  |
|  |  | 19 | 34.0 |  |  |  |
|  |  | 20 | 40.7 |  |  |  |
| 46-50 | M | 2 | 81.4 | Yes | Yes | No |
|  |  | 3 | 74.5 |  |  |  |
|  |  | 4 | 48.7 |  |  |  |
|  |  | 5 | 39.8 |  |  |  |
|  |  | 16 | 34.5 |  |  |  |
|  |  | 17 | 54.9 |  |  |  |
| 36-40 | F | 4 | 40.3 | No | No | No |
|  |  | 11 | 23.6 |  |  |  |
| 46-50 | M | 3 | 30.4 | No | No | No |
|  |  | 8 | 35.6 |  |  |  |
|  |  | 9 | 45.0 |  |  |  |
|  |  | 10 | 39.1 |  |  |  |
|  |  | 12 | >150 |  |  |  |
|  |  | 14 | 84.6 |  |  |  |
| 41-45 | M | 2 | 60.8 | Yes | No | No |
|  |  | 3 | 50.2 |  |  |  |
|  |  | 7 | 13.8 |  |  |  |
| 56-60 | M | 6 | 50.5 | No | No | No |
|  |  | 7 | 29.7 |  |  |  |
| 46-50 | M | 2 | 88.8 | Yes | Yes | Yes |
|  |  | 3 | 86.0 |  |  |  |
| 65-70 | M | 3 | 90.5 | Yes | Yes | Yes |
|  |  | 4 | 86.0 |  |  |  |
| 31-35 | M | 2 | 136.9 | No | No | No |
|  |  | 3 | 5.50 |  |  |  |

\*Patient from outside hospital. Data from this patient was therefore not used in Tables 1 or 2.

